# Views on sharing mental health data for research purposes: A qualitative study with people with mental illness

**DOI:** 10.1101/2022.11.03.22281848

**Authors:** E. Watson, S. Fletcher-Watson, E.J. Kirkham

**Affiliations:** University of Edinburgh Medical School, Edinburgh, UK; Centre for Clinical Brain Sciences, University of Edinburgh, Edinburgh, UK

**Keywords:** mental health, health data, patient perspectives, interview, mental illness, qualitative

## Abstract

**Background:** Improved data sharing could have extensive benefits for mental health research and treatment. However, it is vital that data are shared in a way that aligns with the views of people with mental health conditions. Whilst previous research has examined public views of health data sharing, few studies have focused specifically on people with mental illness.

**Methods:** Semi-structured online interviews were conducted with twelve people with a range mental health conditions, including schizophrenia, anxiety, depression, eating disorder and addiction. Interview questions focussed on the risks and benefits of sharing mental health data, how data should be kept safe, and the sensitivity of different types of data.

**Results:** The overarching themes identified were: benefits of sharing mental health data, concerns about sharing mental health data, safeguards, and data types. There was a high level of support for the use of data sharing to facilitate improved knowledge of and treatment for mental health conditions. Concerns included the potential for misuse of data, such as by insurance companies or employers, and the risk of mental health stigma from researchers and healthcare professionals who accessed the data. There was a focus on appropriate safeguards, such as secure storage access procedures.

**Conclusions:** There was a strong sense across participants that more should be done to combat the suffering caused by mental illness, and that appropriate health data sharing could facilitate this. The mental health research community could build on this generally positive attitude to mental health data sharing by ensuring that they follow rigorous best practice which accounts for the specific concerns of people with mental illness.

---

Large data sets, such as those generated from the routinely-collected data in health records, are becoming increasingly important for public health, contributing to improved service implementation, earlier disease prevention and treatment advances (Bates et al., 2014; Fylan et al., 2019; Kruse et al., 2016). In relation to mental health, routinely-collected data has been used for a range of purposes, including developing more effective ways to identify suicide risk (Simon et al., 2018), examining the effect of neighbourhood regeneration on mental health (White et al., 2017), and identifying the extent to which antipsychotic medication is being prescribed for autistic children (Brophy et al., 2018).

Responsible sharing of health data can overcome some of the limitations of traditional participant-based research models by supporting the collection of larger and more representative population samples (Furimsky et al., 2008; Kirkham et al., 2021; Woodall et al., 2010). This is particularly relevant to mental health research, as traditional research approaches necessarily depend upon recruitment of people who face multiple barriers to participation relating to their time, health, and finances (Kaminsky et al., 2003; Wang et al., 2011).

Whilst routinely-collected data are clearly beneficial for health research, they should only be used in a manner that is supported by those who provide such data, and whose clinical treatment will be shaped by derived insights (Carter et al., 2015). Previous work on this topic has found relatively high levels of public support for health data sharing (73 - 93% positive; Buckley et al., 2011; King et al., 2012; Kirkham et al., 2022; Luchenski et al., 2013). However, this support is somewhat conditional, such that individuals typically want their data to be handled by an organisation they trust and used for the public good rather than for profit (Aitken et al., 2016; Jones et al., 2022). Research also suggests that people’s views vary according to the perceived sensitivity of the data, with mental health data cited as an example of sensitive health data (Grant et al., 2013; Robling et al., 2004).

The extent to which people with mental illness agree with this perception of mental health data as especially sensitive, or how they feel more broadly about sharing mental health data, remains unclear (Shen, Sequeira, et al., 2019). This is because the views of people with mental illness are largely missing from literature on health data sharing (Kirkham et al., 2022; Shen, Sequeira, et al., 2019). For example, none of the 25 studies emerging from Aitken et al.’s (2016) systematic literature review on health data sharing had explicitly recruited people with mental illness.

The small amount of prior research which has focused on the preferences of people with mental illness found that factors which influence willingness to share health data include their prior experiences with health care services (Kirkham et al., 2022; Shen, Bernier, et al., 2019; Soni et al., 2021), stigma and the perceived risk of discrimination (Grando et al., 2017), and, similar to the wider general public, their trust in the organisation accessing the data (Caine & Hanania, 2013). There is also early evidence to suggest that people with mental illness may actually be more willing than those without to share their health data for research purposes (Jones et al., 2022), at least once other factors such as lower overall satisfaction with healthcare have been accounted for (Kirkham et al., 2022). However, this finding is not universal (Soni et al., 2020).

Given that mental health research is currently being held back by concerns around the apparent sensitivity a mental health data (Ford et al., 2021), it is essential that the research community bases its decisions on evidence and not assumptions. Research so far has relied heavily on survey data which rarely provide insights into the details of, and rationale behind, people’s general opinions. Acting to improve use of routine mental health data in research requires an understanding of why people hold certain opinions and what factors inspire trust. Therefore, we conducted semi-structured interviews with 12 people with a range of mental health conditions to investigate what they feel about sharing their health data for research purposes.

## Methods

### Participants

Individuals who took part in a previous survey by our research team (which also focused on the topic of sharing mental health data; Kirkham et al., 2022) were asked to leave their email address if they wanted to take part in an interview. This list of email addresses was used to invite people to take part in the present interview study. Prior to issuing invitations, we used the survey responses to identify potential participants with a diverse range of mental health conditions. We also sought specifically to invite participants from the minority who had responded “no” when the survey asked if they would share their mental health data for research purposes. This was challenging because the vast majority of survey participants (89.7%) had responded “yes” to this question, and those who didn’t often also did not consent to be followed up for this study. Participant demographics, presented in Table 1, were extracted from their aforementioned survey responses. Participants’ experiences of mental illness were extracted from the survey data and confirmed at the beginning of the interview. All participants lived in the UK. The mean age of participants was 40 years, with an age range of 27 to 49 years.

**Table 1:**
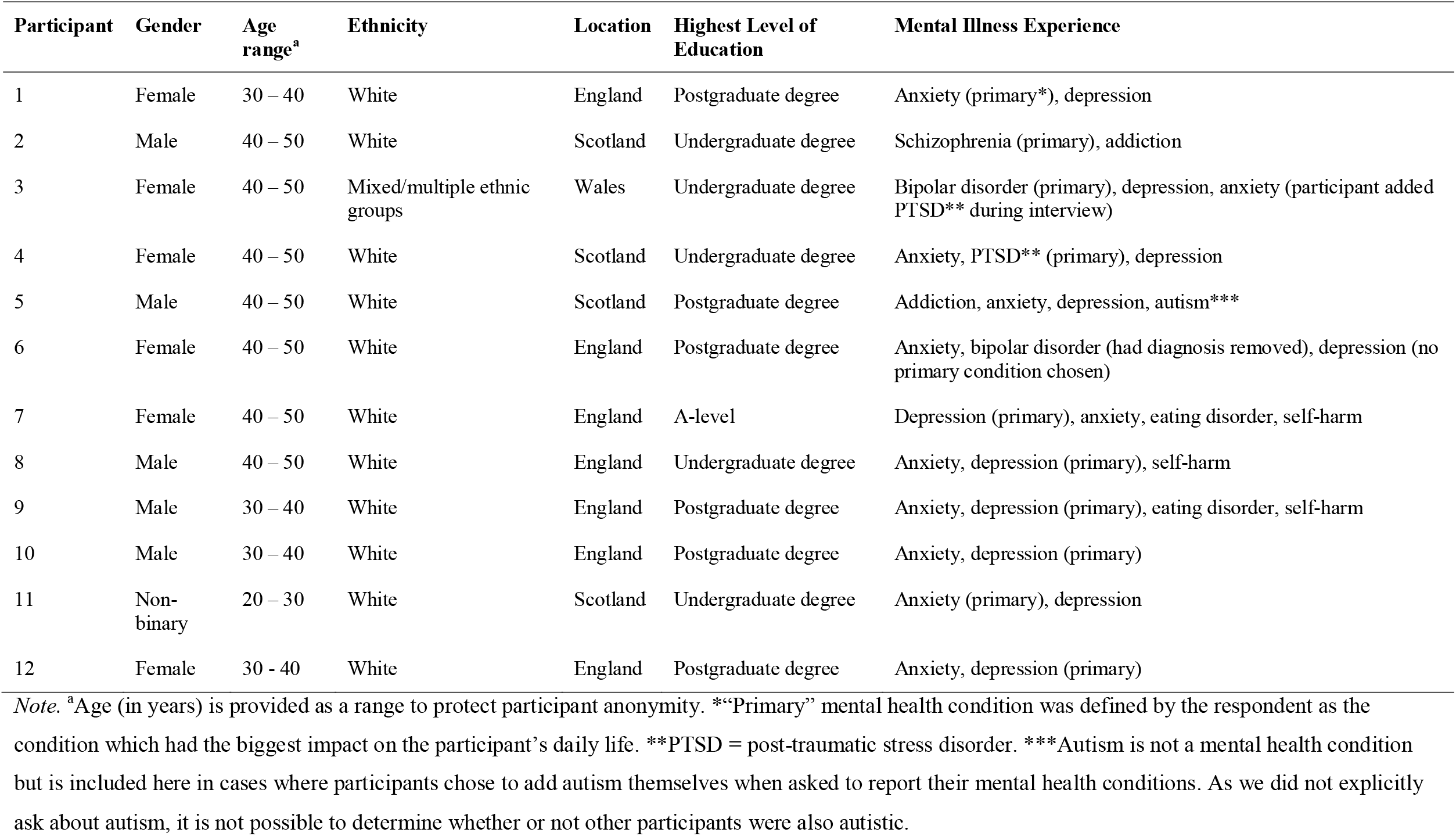
Participant Demographics

### Positionality statement

This paper is written by three white women from the UK, two of whom are researchers and one of whom is a medical student at the time of writing. The interviewer is a female researcher who has lived experience of mental illness, a PhD in Psychology, and professional expertise in working with mental health data. She is part of a university research group which uses Big Data to answer questions about mental health. The person who analysed the data is a female fifth-year medical student (at the time of writing) who has completed a placement in psychiatry. She has previous experience in qualitative research during her intercalated BSc year and a BSc in Physical Activity for Health. The student who completed the analysis was supervised by the researcher who conducted the interviews.

### Interview Guide

The interview guide was developed through discussion within the research team, including an advisory group of community representatives (with lived experience of mental illness and working in mental health services) around the question “How do people feel about data sharing?”. Interview scripts from previous qualitative research were used for guidance. The interview guide was semi structured, and the interviewer asked complementary questions if she deemed it appropriate. The interviewer engaged in a practice interview with Suzy Syrett, a peer researcher who has lived experience of mental illness and expertise in conducting research interviews. The full interview topic guide is available in the Supplementary Materials.

### Procedure

Semi-structured interviews were carried out online via Skype between July and November 2019. The researcher continued to conduct interviews until the point at which perceived data saturation was reached (Morse, 1995; Sandelowski, 1995): i.e. there was the subjective impression that each new participant was largely presenting perspectives already represented in the data. No other individuals were present at the time of interview. Before asking the pre-determined questions, the interviewer allowed time for the participant to become comfortable. It has been shown that allowing for this familiarity to develop fosters in-depth discussions (Braun & Clarke, 2013; Elliott et al., 1999).

Interviews were recorded using the MP3 Skype recorder, which recorded only audio. The researcher who conducted the interviews transcribed five of the interviews and the remainder were transcribed by a paid professional transcriber. Due to a technical error, one interview was not recorded. In this case the interviewer’s notes were used instead. All identifiable information was removed from transcripts.

### Ethics

Ethical approval for this study was provided by the Department of Clinical and Health Psychology Ethics Research Panel, University of Edinburgh, ref STAFF147.

### Data Analysis

The Framework Method was employed to analyse the data. This is an appropriate analysis technique for medical research (Gale et al., 2013). It is a form of thematic analysis not constrained to a specific philosophical or theoretical approach. This enables flexibility in extracting themes and determining parallels and divergences in data (Braun & Clarke, 2013; Gale et al., 2013). Themes were not pre-planned and were derived from the dataset, but they were shaped by the targeted research questions of the research: namely to discover how people with mental illness viewed data sharing. The steps of the Framework Method are outlined in Figure 1. Following analysis, a thematic tree was created.

**Figure 1:**
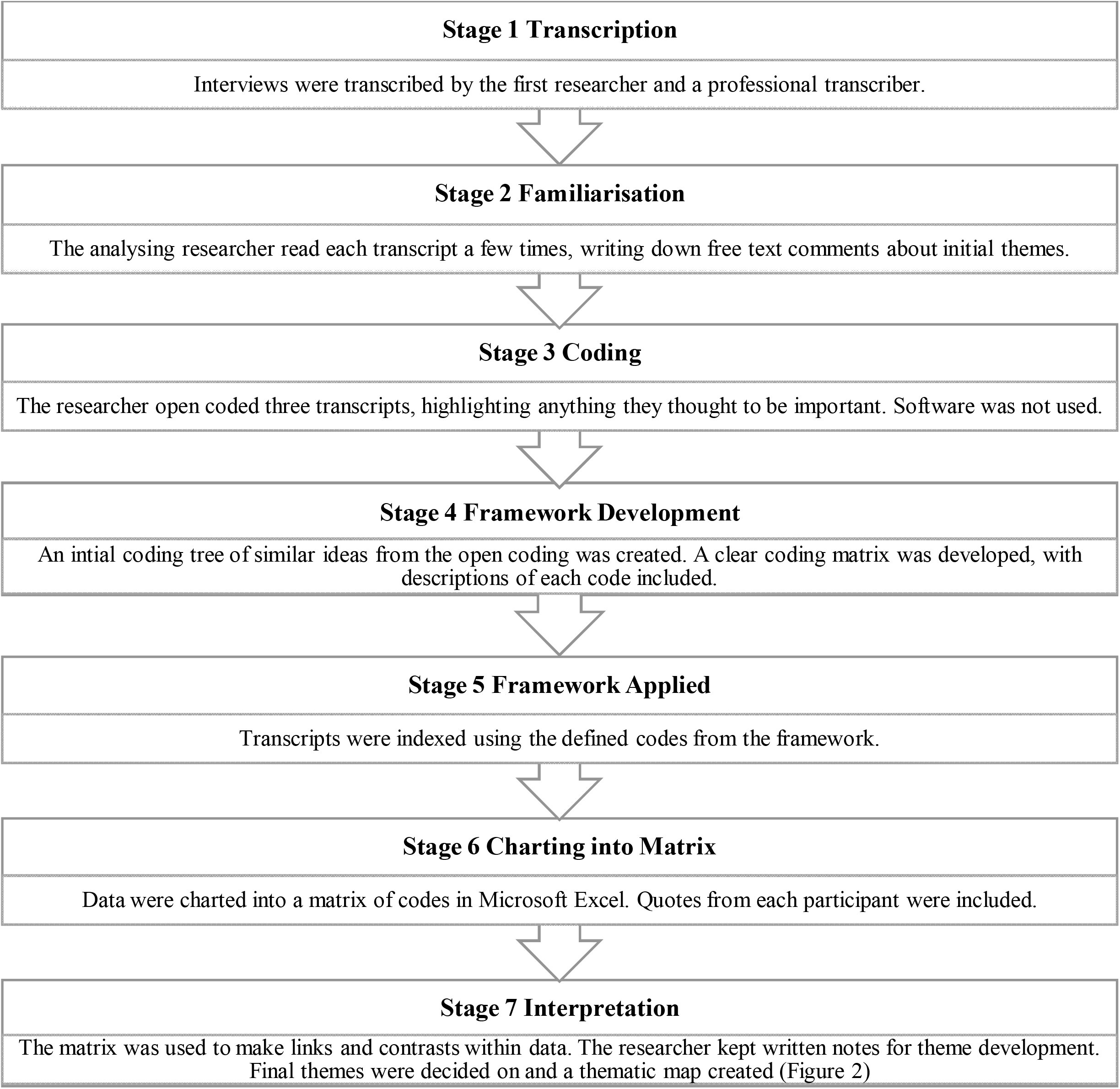
The Framework Method (Amended from Gale et al., 2013)

**Figure 2:**
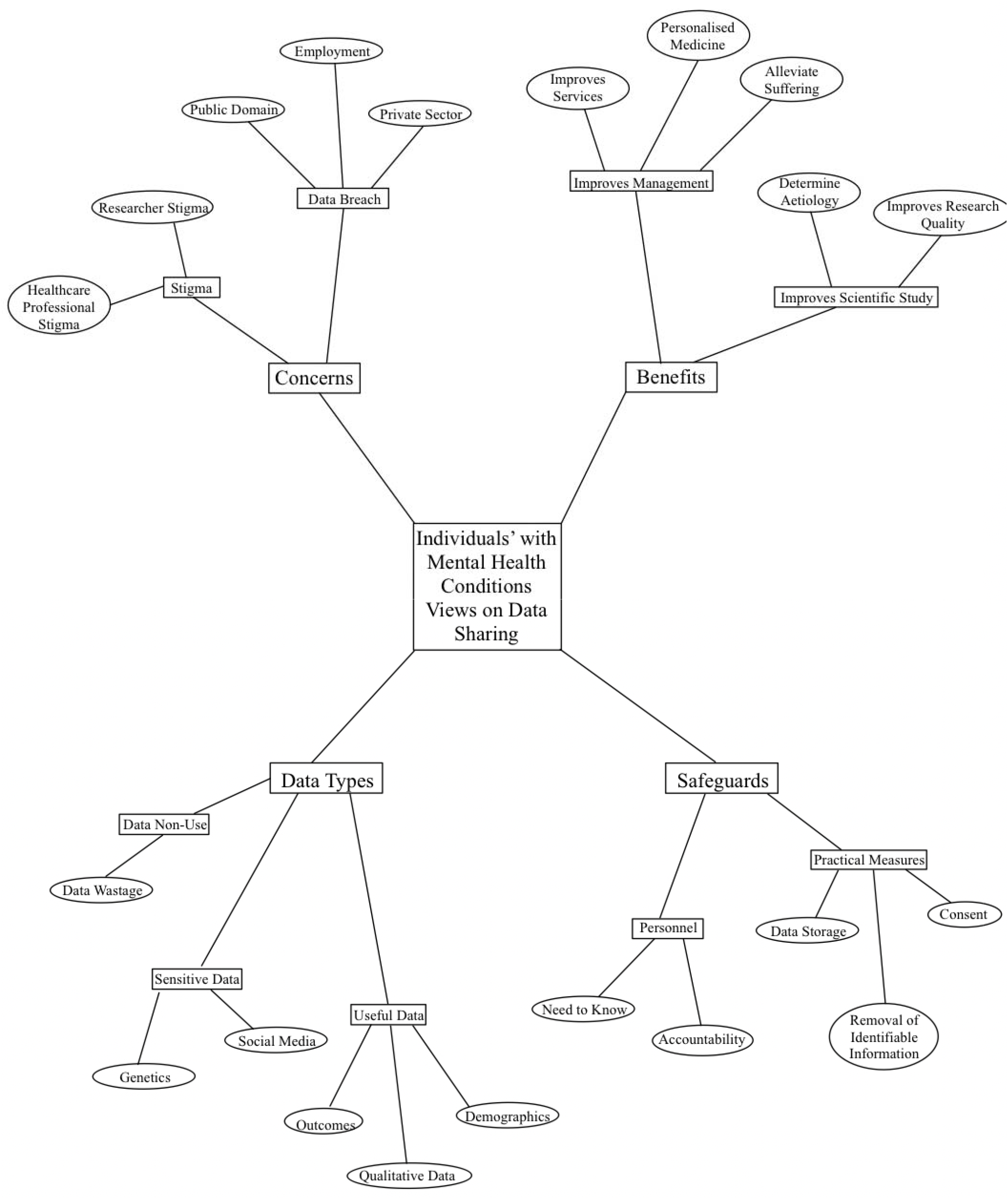
Hierarchical Thematic Map

## Results and Discussion

The aim of this project was to investigate how people with mental illness feel about sharing mental health data for research purposes. Through framework analysis, four top level themes were identified. Nine subthemes within these were identified. The themes identified are illustrated in Table 2, accompanied by an illustrative quote. Most participants were positive about data sharing for mental health research purposes, with one stating *“I am overwhelmingly I think, positive about data sharing, including with mental health*”. In order to avoid repetition, reporting and interpretation of these themes have been included together in one “Results and Discussion” section.

**Table 2:**
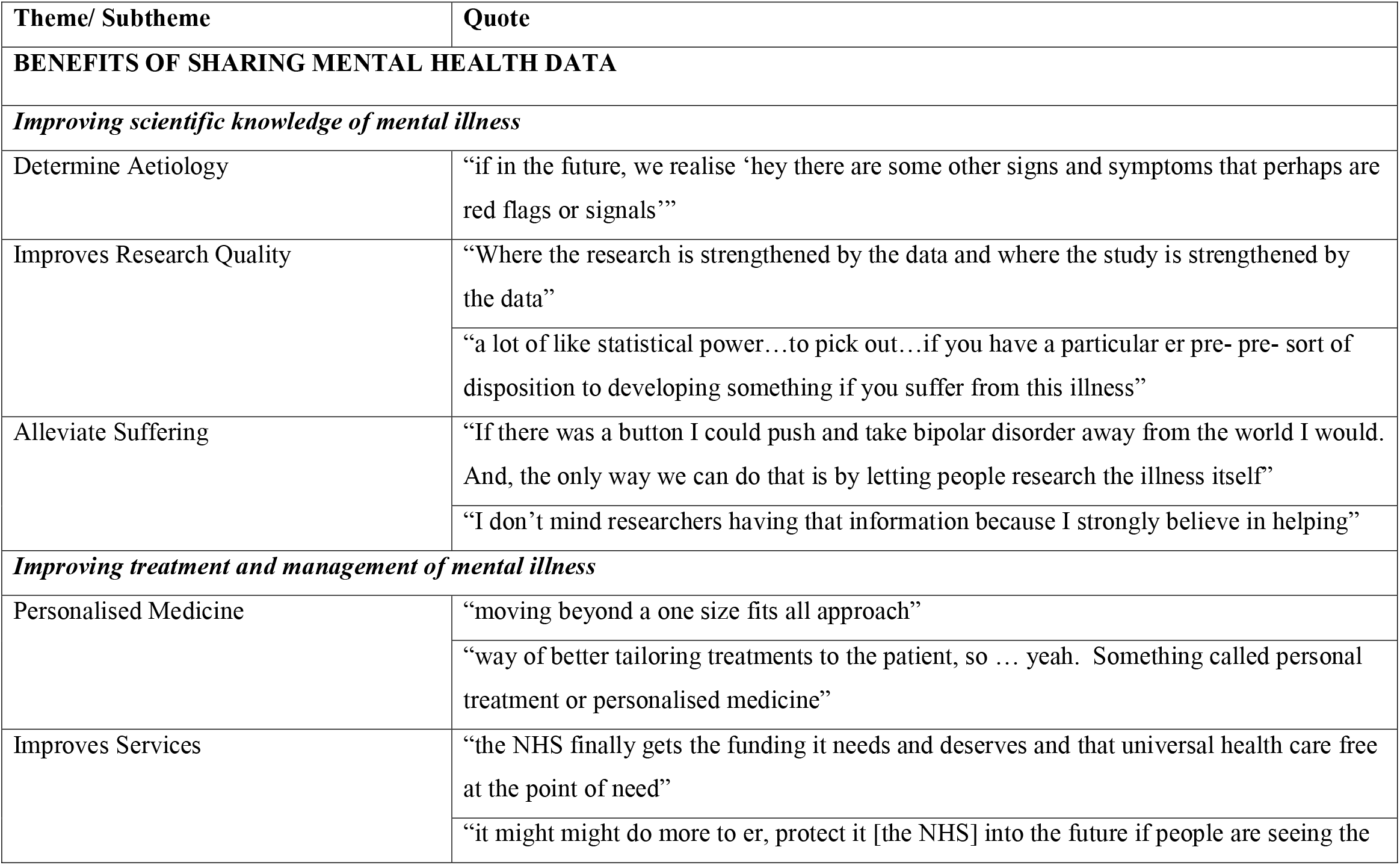
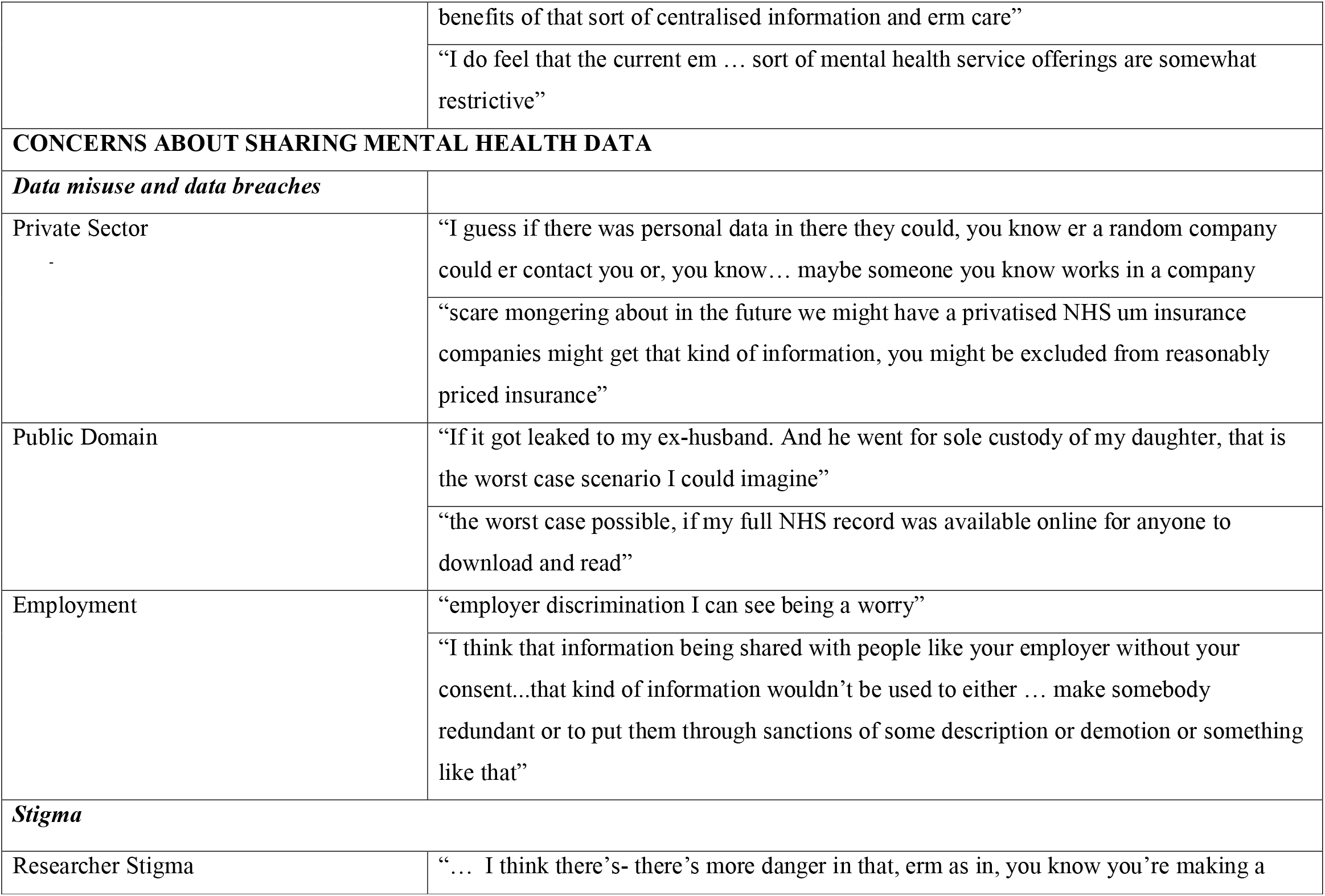
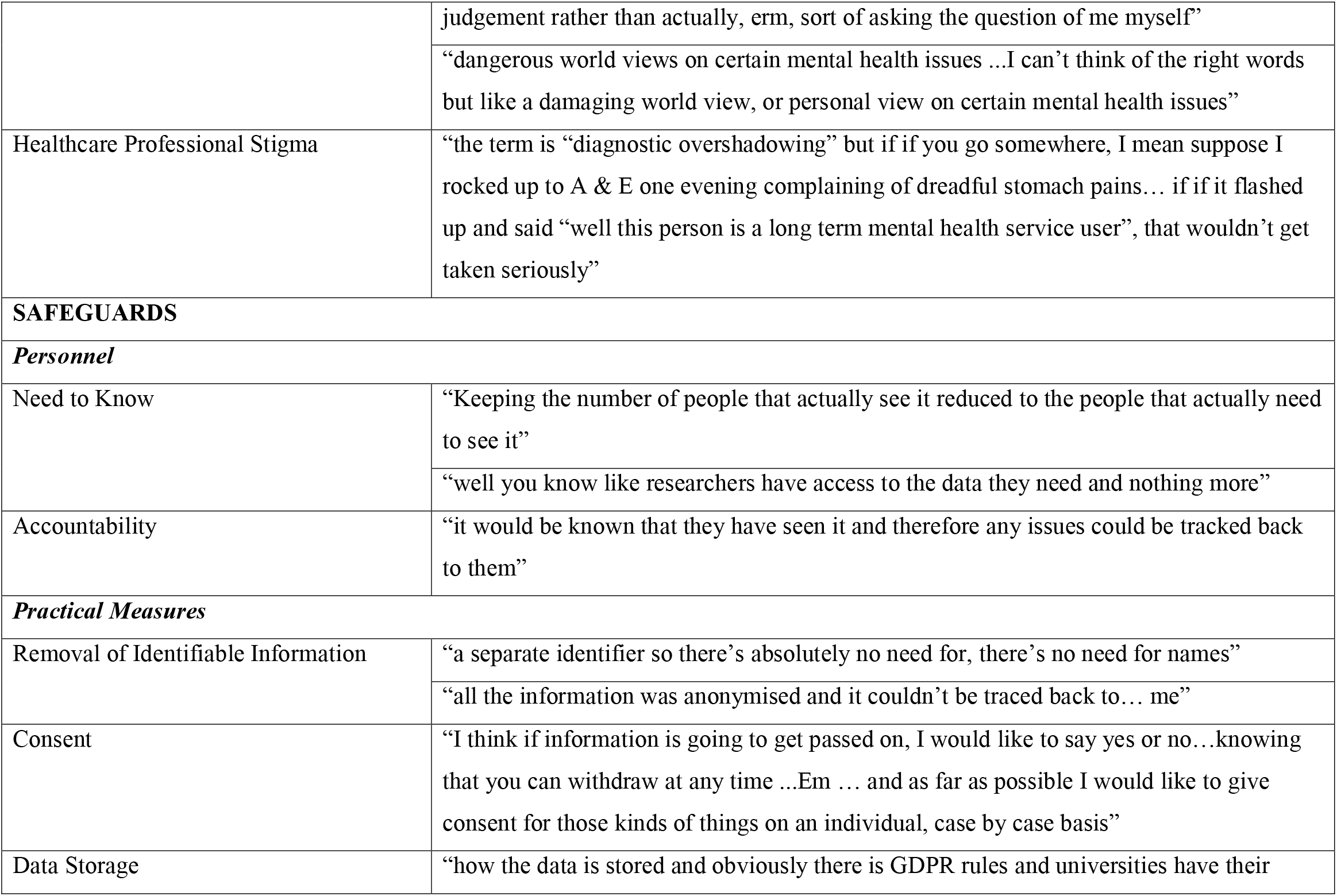
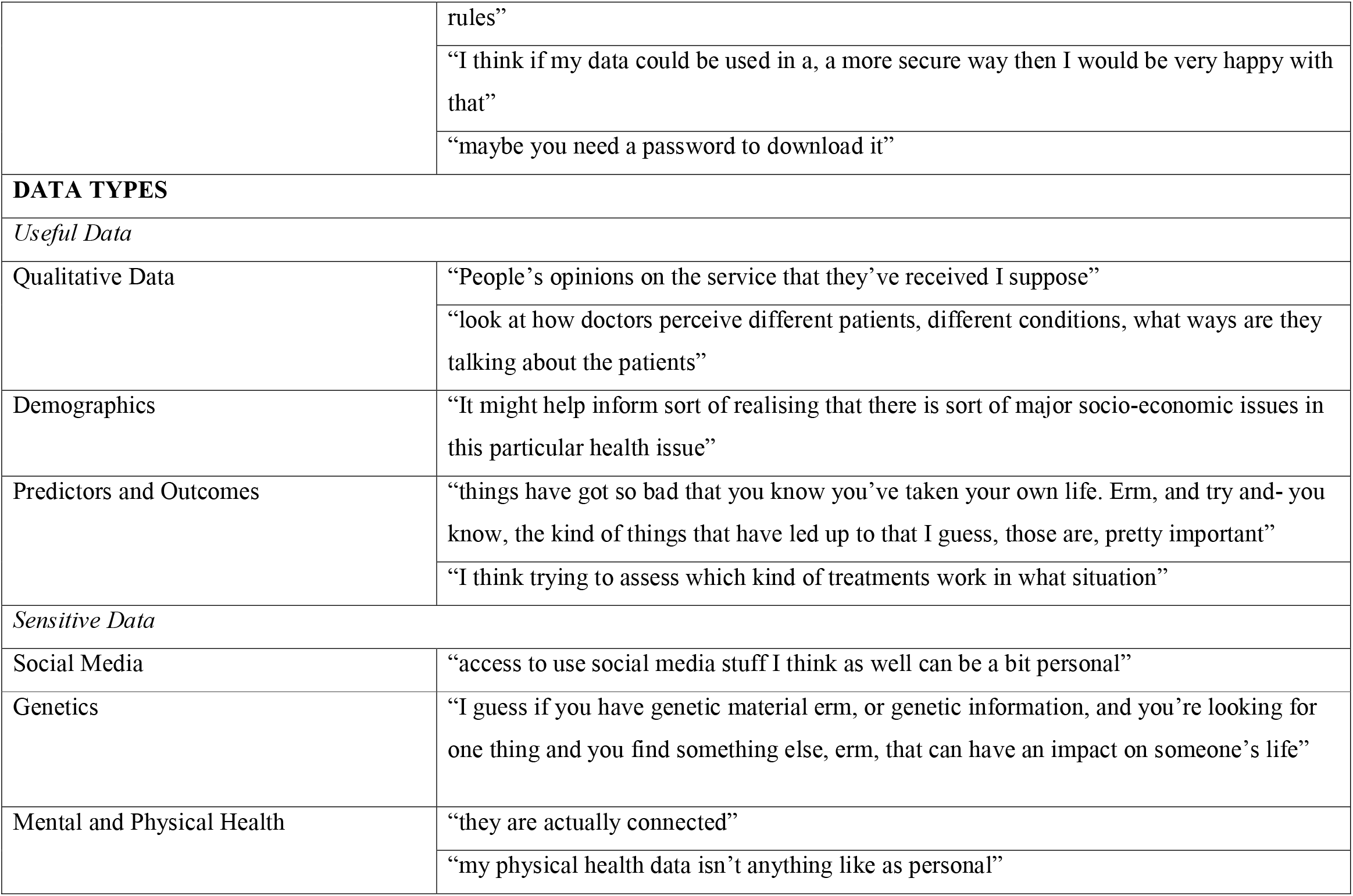
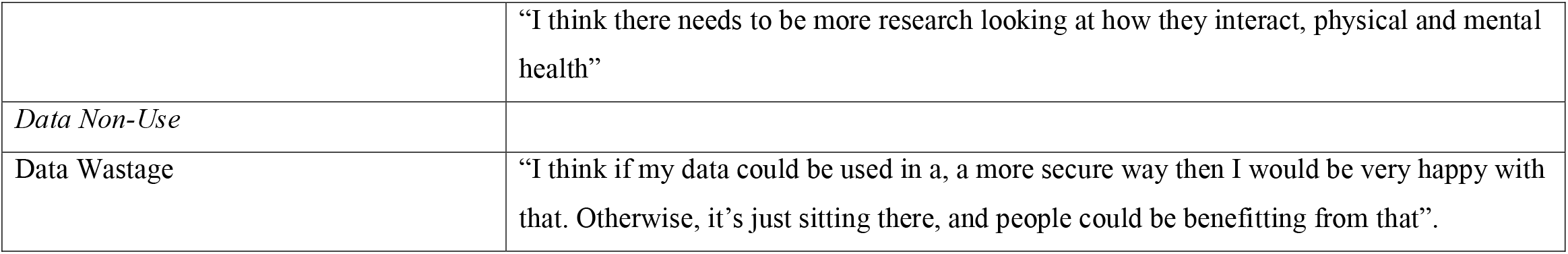
Themes

### Theme 1: Benefits of sharing mental health data

#### Subtheme 1: Improving scientific knowledge of mental illness

Several participants described the potential benefits to scientific research from increased data sharing, using phrases such as “statistical power” in reference to the ability to uncover more effects with larger sample sizes. Participants talked about the role of research in uncovering the aetiology of mental health conditions, and valued the role of data in supporting robust research.

There was an overarching perception that sharing data could help to relieve the suffering of other people with mental health conditions. One participant said, *“if there was a button I could push and take bipolar disorder away from the world I would”*. Further, one participant stated that *“I don’t mind researchers having that information because I strongly believe in helping”*. These findings are very similar to those of a thematic systematic review looking at broader public opinion on data sharing for public health research, which found that a dominant theme was perceived benefits to community, public and science (Howe et al., 2018). Given the strength of feeling from many of the participants in the present research, it is possible that this altruistic approach to sharing health data may be particularly salient amongst people with mental illness. Future work should examine this possibility in further detail.

#### Subtheme 2: Improving treatment and management of mental health conditions

Participants believed that sharing mental health data could improve mental health treatment, with some discussing the concept of *“personalised medicine,”* in which treatments are tailored to a specific patient or group of patients. This focus on personalised medicine may be related to the recent attention to this type of medicine in the physical health sphere (Goetz & Schork, 2018). The adoption of a personalised medicine approach has helped to reduce mortality in cancer and heart disease, yet remains absent from mental health care, in part due to an historical lack of large mental health datasets (Trivedi, 2016). The value of personalised medicine for mental health is highlighted by several of the participants’ own experiences of “trial and error”, with one individual expressing frustration about being placed on eleven different medications before finding one that worked.

A strong focus amongst participants was the potential for data sharing to improve mental health services, with one participant stating they hoped that *“the NHS finally gets the funding it needs”*. A reason for this focus on service improvement may be related to participants’ negative personal experiences of services. Individuals referred to UK mental health services as being “*restrictive*”, “*shocking*” and “*frustrating*”. It is important to note that these comments were made in 2019, prior to the onset of the Covid-19 pandemic. This highlights that whilst the pandemic has increased the burden on mental health services (Lavis, 2022), these services were already struggling to meet people’s needs prior to the pandemic (Kirkham et al., 2022). Interestingly, previous research has shown that positive experiences of mental health services are associated with more positive attitudes to mental health data sharing (Kirkham et al., 2022). Hence, improving people’s experiences of NHS health care could promote data sharing (Fortin et al., 2017; Kirkham et al., 2022)

### Theme 2: Concerns about sharing mental health data

#### Subtheme 3: Data misuse and data breaches

Many participants discussed concerns about data breaches, ranging from data misuse by private sector companies to impacts on their personal life. They expressed fear of being *“excluded from reasonably priced insurance”*, worries about *“employer discrimination”* and even losing child custody. Concern about commercial use of personal health data is strongly reflected in the wider literature on public attitudes to data sharing (Fylan & Fylan, 2021; Howe et al., 2018; Jones et al., 2022; Trinidad et al., 2010).

#### Subtheme 4: Stigma

Concerns about being subject to stigma from the researchers who would potentially have access to their data came up several times, with one participant worrying that researchers may have *“dangerous world views on certain mental health issues*.*”* This finding is supported by previous research citing that the public is less willing to share data with researchers than with healthcare professionals (Fylan & Fylan, 2021). Future interventions focused on improving trust in research institutions may be beneficial, for example ensuring researchers follow best practice guidance when working with mental health data (Kirkham et al., 2021; Kirkham et al., 2020). Though healthcare professionals may be more trusted with mental health data overall, there were nevertheless concerns about stigma from these individuals as well. For example, one participant discussed being wary of increased data sharing due to the concept of *“diagnostic overshadowing”*. This is a phenomenon in which doctors wrongly assume symptoms of physical illness, such as tachycardia, to be a result of co-morbid mental illness (Nash, 2013).

### Theme 3: Safeguards

#### Subtheme 5: Personnel

Participants believed that researchers should have access to their mental health data on a *“need to know”* basis and that they should only be able to access specific information relevant to their research. Further, the concept of researcher accountability and institutional oversight appeared important, with one individual stating they would like any issues to be *“tracked back”* to the researcher. Accountability and oversight were also key elements identified in a recent Delphi study which resulted in a checklist for stakeholder-informed best practice in mental health data science (Kirkham et al. 2021)

#### Subtheme 6: Practical Measures

Overall participants appeared to have a high degree of understanding of safeguards in place for maintaining confidentiality within research. This may be due to the high proportion of participants with postgraduate degrees and relevant professional experience in the current sample. In fact, one individual said, *“I am fairly knowledgeable member of the public, so I don’t know if I am typical in that sense”*. Most participants were happy for their data to be shared if all identifiable information was removed. However, some participants voiced concerns about being identified even if “*pseudonyms*” were used. Therefore, researchers must consider the risk of re-identification from what appears to be fully anonymised data (Ford et al., 2021; Ohm, 2009).

The principle of informed consent seemed very important to participants with regards to data sharing, with one participant saying, “*I think if information is going to get passed on, I would like to say yes or no…knowing that you can withdraw at any time*.*”* This finding echoes a previous focus group study, which looked at the general publics’ opinions about data sharing from primary care medical records. Within this study, individuals voiced anxieties surrounding data collected without consent and felt that the ability to opt-out of data collection was essential (Robling et al., 2004). In terms of the present research, there were inconsistencies between participants with regards to what they considered to be appropriate consent processes. One individual wanted to give consent on a *“case by case”* basis, whilst others didn’t feel as strongly. This contrast is reflected in the literature regarding consent data sharing in terms of general health records (Asai et al., 2002; Cheah et al., 2015; Howe et al., 2018).

The topic of consent is one of the most challenging within health data sharing. Whilst members of the public sometimes express, as above, that they want to provide consent on a case-by-case basis, this is impractical at the level of thousands of individual studies, drawing on data from samples in the millions. Notably, Aitken et al. (2016) found in their systematic review that, though their initial preference may be for opt-in consent, participants typically move away from this model of consent following discussion of its implications. In particular, the need for informed consent would counteract many of the benefits of research with shared routine data also highlighted by our participants, such as the unique opportunities afforded by large sample sizes. Nevertheless, it is important that policy decisions acknowledge the broad range of views surrounding the topic of consent. One suggestion proposed by Jones et al. (2022) would be to manage NHS health data on an opt-out basis, with participants given the option to opt-out on the basis of whether the data would be used for clinical or research purposes, and whether or not it would be potentially identifiable.

### Theme 4: Data Types

#### Subtheme 7: Useful Data

A few participants identified that demographic data about people with mental health conditions might be particularly useful in research, with one individual stating that acquisition of such data *“might help inform sort of realising that there is sort of major socio-economic issues in this particular health issue”*. Furthermore, qualitative data such as *“opinions on the service”* and how doctors are *“talking about the patients”* were deemed particularly relevant to research. Several participants also mentioned that it may be of great value to collect data about which factors can precipitate suicide.

#### Subtheme 8: Sensitive Data

Participants expressed opinions about certain data types being particularly sensitive. Some referred to social media as being *“a bit personal”*. Notably, many participants identified genetic data as being particularly sensitive, which reflects previous research (Trinidad et al., 2010). Their concerns included the potential for “*eugenics*”. As aforementioned, identification with regards to mental health data worried participants.

Participants held contrasting views about the relative sensitivities of physical and mental health data. Some participants believed that they should be considered “*equal*”, whilst others made it clear they thought that mental health data was more personal. This suggests that the wider public perception of mental health data being more sensitive is at least in part shared by people with mental health conditions (King et al., 2012; Robling et al., 2004; Trinidad et al., 2010).

#### Subtheme 9: Data wastage

On a positive note, one participant said *“I think if my data could be used in a, a more secure way then I would be very happy with that. Otherwise, it’s just sitting there, and people could be benefitting from that”*. This quote draws reference to the concept of ‘harm due to data non-use’. In their case study paper, Jones et al (2017) describe how ‘data non-use’ may be associated with increased risk of patient deaths and financial consequences for the health service. They describe that this ‘data non-use’ can be due to several reasons including stringent use of governance frameworks in research (Ford et al., 2021; Jones et al., 2017).

## Strengths and Limitations

A key strength of this study is that the sample was made up of individuals with mental health conditions, whose opinions are rarely prioritised in research on health data sharing. Importantly, individuals had experience with a variety of mental health conditions, including schizophrenia, bipolar disorder and depression. The results of qualitative research are not designed to be applicable to the entire population. Nonetheless, a limitation of this study is the lack of racial diversity within the dataset. Eleven of the twelve participants were white. As it is known that ethnicity and other minority characteristics are related to mental health (Coid et al., 2008; Iwamasa et al., 2002; Meyer et al., 2014; Williams, 2018), and there are multiple historical examples of racism leading to unethical practice in health research. It is important that future qualitative studies consider the impact of intersectionality on opinions about sharing mental health data. In addition, the current interview sample was highly educated, which may have influenced their knowledge of and attitudes towards data sharing.

It is possible that self-selection bias was present. It is feasible that individuals who volunteered to participate in an interview on the topic of mental health data hold more positive views towards mental health data sharing than those with mental illness who did not volunteer. As described above, participants were invited to take part if they had agreed to be contacted for this purpose following completion of our previous survey on health data sharing (Kirkham et al., 2022). When recruiting for the present study we actively tried to invite individuals who had previously responded in the survey that “no”, they were unwilling to share their mental health data for research purposes. However, given that only 10.3% of the full survey sample selected this option, this was challenging, and as a result only two of the final interview sample had responded “no”. Despite this, the participants did vary on a more nuanced survey question in which they used a 5-point Likert scale to indicate how likely they would be to share mental health data, with four of the 12 participants responding “very unlikely” or “unlikely”. A final limitation is that data saturation was judged by the researcher who conducted the interviews. This can be difficult to establish and is usually limited by researcher experience (Tran et al., 2017). A more objective method to calculate the point of data saturation could have been used (Guest et al., 2020).

## Implications and Future Research

In this study, most participants deemed their mental health to be relatively stable at the time of the interview. As such, future research could look specifically at views of individuals currently experiencing poor mental health, although, recruitment amongst this population would pose its own difficulties (McIntosh et al., 2016). As identified, fear of stigma is a major concern with regards to mental health data sharing. This suggests that attempts to reduce mental health stigma in broader society could facilitate more engagement with data sharing initiatives. Findings from this project suggest that knowledge about safeguards in place to protect their data is important to individuals. As such, researchers should endeavour to make these processes transparent, such as by following published guidance on good practice in mental health data sharing (e.g. Kirkham et al., 2020).

## Conclusion

In this interview study with people with lived experience of mental illness we found overarching positive feelings towards the use of mental health data for research which would benefit future treatment and services for people with mental illness. Individuals expressed concerns about data breaches and mental health stigma. There was a strong emphasis placed on safeguards such as anonymisation, appropriate data storage and informed consent. The findings broadly support researchers’ calls for more streamlined access to mental health data under appropriate conditions (Ford et al., 2021). Going forward, the research community should seek to ensure that policy and infrastructure functions to facilitate mental health data science in a manner that is supported by people living with mental illness (Kirkham et al., 2021).

## Data Availability

Data from the interview transcripts are not available as they contain personal and potentially identifiable information. Furthermore, participants did not provide explicit consent for the data to be released beyond the research team that conducted the project.

## References

Aitken, M., Jorre, J. D., Pagliari, C., Jepson, R., & Cunningham-Burley, S. (2016). Public responses to the sharing and linkage of health data for research purposes: a systematic review and thematic synthesis of qualitative studies. Bmc Medical Ethics, 17, Article 73. https://doi.org/10.1186/s12910-016-0153-x

Asai, A., Ohnishi, M., Nishigaki, E., Sekimoto, M., Fukuhara, S., & Fukui, T. (2002). Attitudes of the Japanese public and doctors towards use of archived information and samples without informed consent: Preliminary findings based on focus group interviews. BMC Medical Ethics, 3(1), 1–10.

Bates, D. W., Saria, S., Ohno-Machado, L., Shah, A., & Escobar, G. (2014). Big Data In Health Care: Using Analytics To Identify And Manage High-Risk And High-Cost Patients. Health Affairs, 33(7), 1123–1131. https://doi.org/10.1377/hlthaff.2014.0041

Braun, V., & Clarke, V. (2013). Successful Qualitative Research: A Practical Guide for Beginners.

Brophy, S., Kennedy, J., Fernandez-Gutierrez, F., John, A., Potter, R., Linehan, C., & Kerr, M. (2018). Characteristics of Children Prescribed Antipsychotics: Analysis of Routinely Collected Data. Journal of Child and Adolescent Psychopharmacology, 28(3), 180–191. https://doi.org/10.1089/cap.2017.0003

Buckley, B. S., Murphy, A. W., & MacFarlane, A. E. (2011). Public attitudes to the use in research of personal health information from general practitioners’ records: a survey of the Irish general public. J Med Ethics, 37(1), 50–55. https://doi.org/10.1136/jme.2010.037903

Caine, K., & Hanania, R. (2013). Patients want granular privacy control over health information in electronic medical records. Journal of the American Medical Informatics Association, 20(1), 7–15. https://doi.org/10.1136/amiajnl-2012-001023

Carter, P., Laurie, G. T., & Dixon-Woods, M. (2015). The social licence for research: why care.data ran into trouble. Journal of Medical Ethics, 41(5), 404–409. https://doi.org/10.1136/medethics-2014-102374

Cheah, P. Y., Tangseefa, D., Somsaman, A., Chunsuttiwat, T., Nosten, F., Day, N. P. J., … Parker, M. (2015). Perceived Benefits, Harms, and Views About How to Share Data Responsibly: A Qualitative Study of Experiences With and Attitudes Toward Data Sharing Among Research Staff and Community Representatives in Thailand. Journal of Empirical Research on Human Research Ethics, 10(3), 278–289. https://doi.org/10.1177/1556264615592388

Coid, J. W., Kirkbride, J. B., Barker, D., Cowden, F., Stamps, R., Yang, M., & Jones, P. B. (2008). Raised incidence rates of all psychoses among migrant groups: findings from the East London first episode psychosis study. Arch Gen Psychiatry, 65(11), 1250–1258. https://doi.org/10.1001/archpsyc.65.11.1250

Elliott, R., Fischer, C. T., & Rennie, D. L. (1999). Evolving guidelines for publication of qualitative research studies in psychology and related fields. Br J Clin Psychol, 38(3), 215–229. https://doi.org/10.1348/014466599162782

Ford, T., Mansfield, K. L., Markham, S., McManus, S., John, A., O’Reilly, D., … Shenow, S. (2021). The challenges and opportunities of mental health data sharing in the UK. Lancet Digit Health, 3(6), e333–e336. https://doi.org/10.1016/s2589-7500(21)00078-9

Fortin, M., Bamvita, J.-M., & Fleury, M.-J. (2017). Patient satisfaction with mental health services based on Andersen’s Behavioral Model. The Canadian Journal of Psychiatry, 63(2), 103–114. https://doi.org/10.1177/0706743717737030

Furimsky, I., Cheung, A. H., Dewa, C. S., & Zipursky, R. B. (2008). Strategies to enhance patient recruitment and retention in research involving patients with a first episode of mental illness [Article]. Contemporary Clinical Trials, 29(6), 862–866. https://doi.org/10.1016/j.cct.2008.07.005

Fylan, B., Marques, I., Ismail, H., Breen, L., Gardner, P., Armitage, G., & Blenkinsopp, A. (2019). Gaps, traps, bridges and props: a mixed-methods study of resilience in the medicines management system for patients with heart failure at hospital discharge. BMJ open, 9(2), e023440.

Fylan, F., & Fylan, B. (2021). Co-creating social licence for sharing health and care data. Int J Med Inform, 149, 104439. https://doi.org/10.1016/j.ijmedinf.2021.104439

Gale, N. K., Heath, G., Cameron, E., Rashid, S., & Redwood, S. (2013). Using the framework method for the analysis of qualitative data in multi-disciplinary health research. BMC Med Res Methodol, 13, 117. https://doi.org/10.1186/1471-2288-13-117

Goetz, L. H., & Schork, N. J. (2018). Personalized medicine: motivation, challenges, and progress. Fertil Steril, 109(6), 952–963. https://doi.org/10.1016/j.fertnstert.2018.05.006

Grando, M. A., Murcko, A., Mahankali, S., Saks, M., Zent, M., Chern, D., … Hassanzadeh, N. (2017). A Study to Elicit Behavioral Health Patients’ and Providers’ Opinions on Health Records Consent. Journal of Law Medicine & Ethics, 45(2), 238–259. https://doi.org/10.1177/1073110517720653

Grant, A., Ure, J., Nicolson, D. J., Hanley, J., Sheikh, A., McKinstry, B., & Sullivan, F. (2013). Acceptability and perceived barriers and facilitators to creating a national research register to enable ‘direct to patient’ enrolment into research: the Scottish Health Research Register (SHARE). Bmc Health Services Research, 13, Article 422. https://doi.org/10.1186/1472-6963-13-422

Guest, G., Namey, E., & Chen, M. (2020). A simple method to assess and report thematic saturation in qualitative research. PloS one, 15(5), e0232076–e0232076. https://doi.org/10.1371/journal.pone.0232076

Howe, N., Giles, E., Newbury-Birch, D., & McColl, E. (2018). Systematic review of participants’ attitudes towards data sharing: a thematic synthesis. J Health Serv Res Policy, 23(2), 123–133. https://doi.org/10.1177/1355819617751555

Iwamasa, G. Y., Sorocco, K. H., & Koonce, D. A. (2002). Ethnicity and clinical psychology: a content analysis of the literature. Clin Psychol Rev, 22(6), 931–944. https://doi.org/10.1016/s0272-7358(02)00147-2

Jones, K. H., Laurie, G., Stevens, L., Dobbs, C., Ford, D. V., & Lea, N. (2017). The other side of the coin: Harm due to the non-use of health-related data. International Journal of Medical Informatics, 97, 43–51. https://doi.org/https://doi.org/10.1016/j.ijmedinf.2016.09.010

Jones, L. A., Nelder, J. R., Fryer, J. M., Alsop, P. H., Geary, M. R., Prince, M., & Cardinal, R. N. (2022). Public opinion on sharing data from health services for clinical and research purposes without explicit consent: an anonymous online survey in the UK. Bmj Open, 12(4), Article e057579. https://doi.org/10.1136/bmjopen-2021-057579

Kaminsky, A., Roberts, L. W., & Brody, J. L. (2003). Influences upon willingness to participate in schizophrenia research: An analysis of narrative data from 63 people with schizophrenia. Ethics & Behavior, 13(3), 279–302. https://doi.org/10.1207/s15327019eb1303_06

King, T., Brankovic, L., & Gillard, P. (2012). Perspectives of Australian adults about protecting the privacy of their health information in statistical databases. International Journal of Medical Informatics, 81(4), 279–289. https://doi.org/https://doi.org/10.1016/j.ijmedinf.2012.01.005

Kirkham, E. J., Crompton, C. J., Iveson, M. H., Beange, I., McIntosh, A. M., & Fletcher-Watson, S. (2021). Co-development of a Best Practice Checklist for Mental Health Data Science: A Delphi Study [Original Research]. Frontiers in Psychiatry, 12(901). https://doi.org/10.3389/fpsyt.2021.643914

Kirkham, E. J., Iveson, M., Beange, I., Crompton, C. J., McIntosh, A., & Fletcher-Watson, S. (2020). A stakeholder-derived best practice checklist for mental health data science in the UK. In. Edinburgh, UK: University of Edinburgh.

Kirkham, E. J., Lawrie, S. M., Crompton, C. J., Iveson, M. H., Jenkins, N. D., Goerdten, J., … Fletcher-Watson, S. (2022). Experience of clinical services shapes attitudes to mental health data sharing: findings from a UK-wide survey. BMC Public Health, 22(1), 357. https://doi.org/10.1186/s12889-022-12694-z

Kruse, C. S., Goswamy, R., Raval, Y., & Marawi, S. (2016). Challenges and Opportunities of Big Data in Health Care: A Systematic Review. JMIR Med Inform, 4(4), e38. https://doi.org/10.2196/medinform.5359

Lavis, P. (2022). Running hot: the impact of the pandemic on mental health services. N. confederation. https://www.scie-socialcareonline.org.uk/running-hot-the-impact-of-the-pandemic-on-mental-health-services/r/a116f00000Um3A3AAJ

Luchenski, S. A., Reed, J. E., Marston, C., Papoutsi, C., Majeed, A., & Bell, D. (2013). Patient and Public Views on Electronic Health Records and Their Uses in the United Kingdom: Cross-Sectional Survey. Journal of Medical Internet Research, 15(8), Article UNSP e160. https://doi.org/10.2196/jmir.2701

McIntosh, A. M., Stewart, R., John, A., Smith, D. J., Davis, K., Sudlow, C., … Porteous, D. J. (2016). Data science for mental health: a UK perspective on a global challenge. Lancet Psychiatry, 3(10), 993–998. https://doi.org/10.1016/s2215-0366(16)30089-x

Meyer, O. L., Castro-Schilo, L., & Aguilar-Gaxiola, S. (2014). Determinants of mental health and self-rated health: a model of socioeconomic status, neighborhood safety, and physical activity. Am J Public Health, 104(9), 1734–1741. https://doi.org/10.2105/ajph.2014.302003

Morse, J. M. (1995). The Significance of Saturation. Qualitative Health Research, 5(2), 147–149. https://doi.org/10.1177/104973239500500201

Nash, M. (2013). Diagnostic overshadowing: A potential barrier to physical health care for mental health service users. Mental Health Practice, 17, 22–26. https://doi.org/10.7748/mhp2013.12.17.4.22.e862

Ohm, P. (2009). Broken promises of privacy: Responding to the surprising failure of anonymization. UCLA l. Rev., 57, 1701.

Robling, M. R., Hood, K., Houston, H., Pill, R., Fay, J., & Evans, H. M. (2004). Public attitudes towards the use of primary care patient record data in medical research without consent: a qualitative study. Journal of Medical Ethics, 30(1), 104–109. https://doi.org/10.1136/jme.2003.005157

Sandelowski, M. (1995). Sample size in qualitative research. Research in Nursing & Health, 18(2), 179–183. https://doi.org/https://doi.org/10.1002/nur.4770180211

Shen, N., Bernier, T., Sequeira, L., Strauss, J., Silver, M. P., Carter-Langford, A., & Wiljer, D. (2019). Understanding the patient privacy perspective on health information exchange: A systematic review. Int J Med Inform, 125, 1–12. https://doi.org/10.1016/j.ijmedinf.2019.01.014

Shen, N., Sequeira, L., Silver, M. P., Carter-Langford, A., Strauss, J., & Wiljer, D. (2019). Patient Privacy Perspectives on Health Information Exchange in a Mental Health Context: Qualitative Study. Jmir Mental Health, 6(11), Article UNSP e13306. https://doi.org/10.2196/13306

Simon, G. E., Johnson, E., Lawrence, J. M., Rossom, R. C., Ahmedani, B., Lynch, F. L., … Shortreed, S. M. (2018). Predicting Suicide Attempts and Suicide Deaths Following Outpatient Visits Using Electronic Health Records. American Journal of Psychiatry, 175(10), 951–960. https://doi.org/10.1176/appi.ajp.2018.17101167

Soni, H., Grando, A., Murcko, A., Diaz, S., Mukundan, M., Idouraine, N., … Whitfield, M. J. (2020). State of the art and a mixed-method personalized approach to assess patient perceptions on medical record sharing and sensitivity. Journal of Biomedical Informatics, 101, Article 103338. https://doi.org/10.1016/j.jbi.2019.103338

Soni, H., Ivanova, J., Grando, A., Murcko, A., Chern, D., Dye, C., & Whitfield, M. J. (2021). A pilot comparison of medical records sensitivity perspectives of patients with behavioral health conditions and healthcare providers. Health Informatics Journal, 27(2), Article 14604582211009925. https://doi.org/10.1177/14604582211009925

Tran, V. T., Porcher, R., Tran, V. C., & Ravaud, P. (2017). Predicting data saturation in qualitative surveys with mathematical models from ecological research. J Clin Epidemiol, 82, 71-78.e72. https://doi.org/10.1016/j.jclinepi.2016.10.001

Trinidad, S. B., Fullerton, S. M., Bares, J. M., Jarvik, G. P., Larson, E. B., & Burke, W. (2010). Genomic research and wide data sharing: Views of prospective participants. Genetics in Medicine, 12(8), 486–495. https://doi.org/https://doi.org/10.1097/GIM.0b013e3181e38f9e

Trivedi, M. H. (2016). Right patient, right treatment, right time: biosignatures and precision medicine in depression. World Psychiatry, 15(3), 237–238. https://doi.org/https://doi.org/10.1002/wps.20371

Wang, J. L., Patten, S., Currie, S., Sareen, J., & Schmitz, N. (2011). Perceived Needs for and Use of Workplace Accommodations by Individuals With a Depressive and/or Anxiety Disorder. Journal of Occupational and Environmental Medicine, 53(11), 1268–1272. https://doi.org/10.1097/JOM.0b013e31822cfd82

White, J., Greene, G., Farewell, D., Dunstan, F., Rodgers, S., Lyons, R. A., … Fone, D. (2017). Improving Mental Health Through the Regeneration of Deprived Neighborhoods: A Natural Experiment. American Journal of Epidemiology, 186(4), 473–480. https://doi.org/10.1093/aje/kwx086

Williams, D. R. (2018). Stress and the Mental Health of Populations of Color: Advancing Our Understanding of Race-related Stressors. Journal of health and social behavior, 59(4), 466–485. https://doi.org/10.1177/0022146518814251

Woodall, A., Morgan, C., Sloan, C., & Howard, L. (2010). Barriers to participation in mental health research: are there specific gender, ethnicity and age related barriers? [Article]. Bmc Psychiatry, 10, 10, Article 103. https://doi.org/10.1186/1471-244x-10-103

